# Performance of polygenic risk scores in screening, prediction, and risk stratification

**DOI:** 10.1101/2022.02.18.22271049

**Authors:** AD. Hingorani, J. Gratton, C. Finan, AF. Schmidt, R. Patel, R. Sofat, V. Kuan, C. Langenberg, H. Hemingway, JK. Morris, NJ. Wald

## Abstract

**Background:** The clinical value of polygenic risk scores has been questioned. We sought to clarify performance in population screening, individual risk prediction and population risk stratification by analysing 926 polygenic risk scores for 310 diseases from the Polygenic Score (PGS) Catalog.

**Methods:** Polygenic risk scores in the PGS Catalog are reported using hazard ratios or odds ratios per standard deviation, or the area under the receiver operating characteristic curve sometimes expressed as the *C*-index. We used this information to produce estimates of performance in: (a) *population screening* — by calculating the detection rate (*DR*_5_) for a 5% false positive rate (*FPR*) and the population odds of becoming affected given a positive result (*OAPR*); (b) *individual risk prediction* — by calculating the individual odds of becoming affected for a person with a particular polygenic score; and (c) *population risk stratification* — by calculating the odds of becoming affected for groups of individuals in different portions of a polygenic risk score distribution. We use coronary artery disease and breast cancer as illustrative examples.

**Findings:** *Population screening performance*: The median *DR*_5_ for all polygenic risk scores and all diseases studied was 11% [interquartile range 8 − 18%]. The median *DR*_5_ was 12% [9 − 19] for polygenic risk scores for CAD and 10% [9 − 12] for breast cancer, with population *OAPRs* of 1:8 and 1: 21 respectively, with background 10-year odds of 1:19 and 1:41 respectively, which are typical for these diseases at age 50. *Individual risk prediction*: The corresponding 10-year odds of becoming affected for individuals aged 50 with a polygenic risk score at the 2.5*th*, 25*th*, 75*th* and 97.5*th* centile were 1:54, 1:29, 1:15, and 1:8 for CAD and 1:91, 1:56, 1:34, and 1:21 for breast cancer. *Population risk stratification*: At age 50, stratifying into quintile groups of CAD risk yielded 10-year odds of 1: 41 and 1: 11 for the lowest and highest quintile groups respectively. The 10-year odds was 1: 7 for the upper 2.5% of the polygenic risk score distribution for CAD, a group that contributed 7% of cases. The corresponding estimates for breast cancer were 1: 72 and 1: 26 for lowest and highest quintiles; and 1:19 for the upper 2.5% of the distribution, which contributed 6% of cases.

**Interpretation:** Polygenic risk scores perform poorly in population screening, individual risk prediction, and population risk stratification.

**Funding:** British Heart Foundation; UK Research and Innovation; National Institute of Health and Care Research.

## Introduction

A polygenic risk score represents the weighted sum of independent DNA sequence variants that are present in an individual’s genome which increase the risk of a particular disease^1^. The weight assigned to each variant is based on the strength of its disease association estimated from a genome wide association study (GWAS). The increasing range and scale of GWAS over the last decade, now spanning over 2500 diseases or traits^2^, has led to a proliferation in polygenic risk scores, igniting widespread interest in potential healthcare applications^3^, and capturing the attention of policy makers^4^.

Claims have been made that polygenic risk scores generate ‘substantial’ improvements in risk prediction^5^, will ‘power a transformative change to healthcare’^6^, and are ready to implement in practice^7^. In a progression toward clinical implementation, position papers have appeared on reporting standards and responsible clinical use from the Clinical Genome Resource (ClinGen) Complex Disease Working Group^8^, and the Polygenic Risk Score Task Force of the International Common Disease Alliance^9^. Individual consumers and healthcare providers can already access commercial genetic testing and software services based on polygenic scores^10–12^. A ‘world-first pilot’ trial of predictive genetic testing for cardiovascular disease is also underway in participants attending Vascular Health Checks in the NHS^13^, and ‘genetic risk scores’ for disease prediction are central to aims of the Our Future Health programme aiming to recruit 5 million UK adults^14^.

These claims, however, are disputed. There is disagreement on the performance of polygenic risk scores in population screening, individual risk prediction and population risk stratification which makes their eventual role in medicine and public health uncertain^15–18^. Recently, Lambert and colleagues produced the PGS Catalog, a comprehensive, regularly updated, open-access directory of studies on *polygenic scores* for quantitative traits (e.g. blood pressure) and *polygenic risk scores* for diseases (e.g. breast cancer)^19^. The catalogue lists the following ‘performance metrics’ for polygenic risk scores: the hazard (*HR*) or odds ratio (*OR*), both per 1-standard deviation (*SD*) increment in the score, or the area under the receiver operating characteristic curve (*AUC*), sometimes expressed as the *C*-index. However, these metrics are not directly informative of performance in population screening, individual risk prediction, or population risk stratification. An appropriate metric is the odds of becoming affected, which is the positive predictive value expressed as an odds. For example, an odds of 1: 9 equates to a risk of 1 in 10 or 10%. The odds of becoming affected is obtained by multiplying the background odds of developing disease in a specified time frame by the likelihood ratio (*LR*) associated with a ‘positive test’ (in the case of population screening), with a particular polygenic score value (in the case of individual risk prediction), or occupancy of a particular polygenic risk score quantile group (in the case of population risk stratification).

We mathematically derive the odds of becoming affected in each of these scenarios using the metrics reported in the PGS Catalog. We use breast cancer and coronary artery disease as illustrative examples and scrutinise two proposed early clinical uses of polygenic risk scores: to to improve on the performance of the established risk factor models in the prediction of CAD and stroke, and to prioritise mammographic screening at a younger age for the detection of breast cancer.

## Methods

By April 2022, the PGS Catalog had curated 13 828 performance metric estimates for 2194 polygenic scores (‘unique polygenic score codes’), involving 544 diseases or traits (‘unique experimental factor ontology identifiers’), reported in 303 unique publications. We removed polygenic scores for continuous traits, and polygenic risk scores with implausible values (167 instances where the *HR* or *OR* per-*SD* was recorded as < 1, two instances where the *AUC* was < 0.5, and one instance where the *C*-index was recorded as 632), leaving 3915 performance metric estimates for 926 polygenic risk scores involving 310 unique binary outcomes (mainly diseases). The reported performance metrics were *OR* per-*SD* in 1216 instances, *HR* per-*SD* in 378 instances, *AUC* in 2077 and *C*-index in 244 instances (**Supplementary Tables)**.

Polygenic risk scores display a Gaussian distribution, so it is possible to use the originally reported metrics in the PGS Catalog to calculate the difference in mean values between individuals who are later affected or remain unaffected by disease (**Supplementary Tables**).^20,21^ We used the difference in mean values to estimate the overlap in Gaussian distributions and calculate the *DR* and *FPR* which are the percentage of people with a polygenic score above a particular cut-off (‘a positive test’) among those who are later affected or remain unaffected by disease, respectively. For simplicity and consistency, we set polygenic score cut-offs that define a 5% *FPR* and calculate the corresponding detection rate (*DR*_5_) ^20,22^. However, we also provide estimates for *DR*_10_ and *DR*_1_, the detection rates at 10% and 1% false positive rates (**Supplementary Tables**). Detection rates at any other false positive rate can be derived by adapting the equations in the **Extended Methods**. We defined the *LR* in screening as the ratio *DR*/*FPR*. In individual risk prediction, we defined *LR* as the ratio of the heights of the Gaussian distribution curves for affected and unaffected individuals at a particular polygenic risk score centile; and in risk stratification as the ratio of areas under the relative frequency distributions for affected and unaffected individuals in each polygenic score quantile (e.g., each fifth, or the upper 2.5% of the polygenic score distribution, see **Extended Methods**). In each case, multiplying the *LR* by the background odds of disease for the whole population gives the corresponding odds of becoming affected for the individual or group of interest. Where we refer to a polygenic score centile or quantile, this is in reference to the distribution in the unaffected group. When referring to a particular polygenic risk score, we used the PGS Catalog identifier number.

We re-analysed data from two original sources^5,23^ to quantify the extent to which the addition of polygenic risk score information to conventional risk factors improves the prediction of CAD and stroke. To do this, we calculated the *DR* and *FPR* with and without polygenic risk score information, using 10-year risk cut-offs recommended in guidelines for the initiation of statin treatment. We then calculated the number of individuals who need to be genotyped (and a polygenic score calculated) to detect or prevent one additional CAD event or stroke. We refer to this value as the ‘number-needed-to-genotype’. We modelled the use of a breast cancer polygenic risk score to prioritise mammographic screening at age 40 rather than from the currently recommended 50 years of age.

## Results

### Performance of polygenic risk scores in screening

For all diseases studied, the median *DR*_5_ based on all polygenic risk scores was 11%, interquartile range [8 − 18%], i.e., 89% [82 − 92] of cases missed. The median *DR*_5_ values for polygenic risk scores whose performance was reported using *OR* or *HR* per-*SD* were 9% [6 − 12] and 8% [7 − 10] respectively. For polygenic risk score performance reported using *AUC* or *C*-index, the median *DR*_5_ values were 14% [10 − 22] and 19% [13 − 25] respectively. The median *DR*_5_ values for polygenic risk scores for 28 common diseases, including CAD and breast cancer are shown in **Figure 1**.

**Figure 1.**
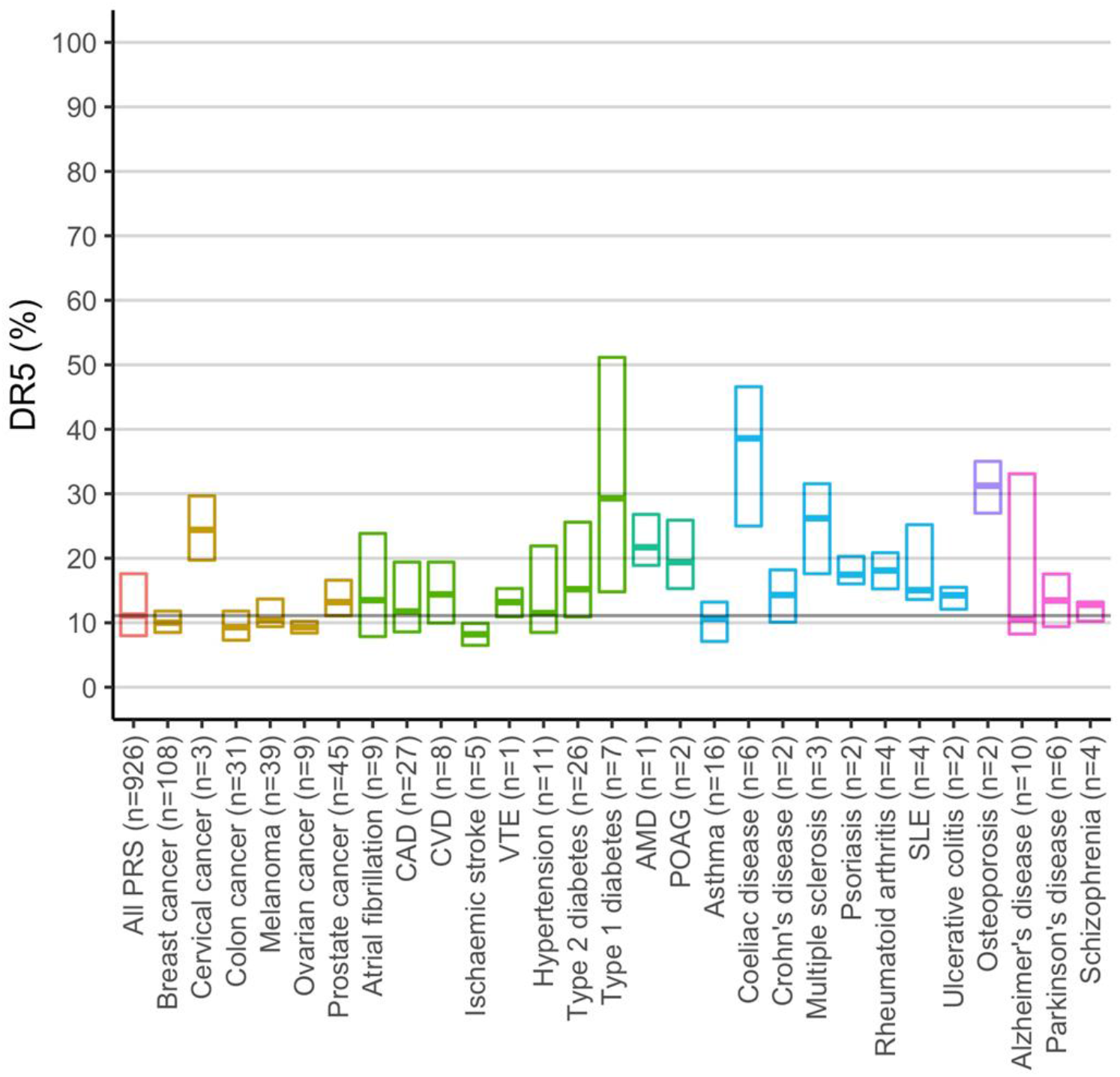
Performance in screening estimated for polygenic risk scores included in the PGS Catalog as of April 2022. The limits of each box represent the interquartile range and the horizontal line within each box is the estimated *DR*_5_ value based on performance metrics reported for the corresponding polygenic risk scores. The selected diseases are colour coded into the following categories: cancers, cardiometabolic conditions, ocular diseases, allergic or autoimmune diseases, bone disease and neuropsychiatric diseases. The horizontal grey line is the estimated median *DR*_5_ value based on performance metrics for all 926 polygenic risk scores and all diseases studied in the PGS Catalog. Abbreviations: PRS – polygenic risk score; CVD – cardiovascular disease; VTE – venous thromboembolic disease; AMD – age-related macular degeneration; POAG – primary open angle glaucoma; SLE – systemic lupus erythematosus.

#### Coronary artery disease

The median *DR*_5_ from performance metrics for 27 polygenic risk scores for CAD was 12% [9 − 20] (88% of cases missed, **Figure 2**), which corresponds to an *LR* of 2.4. Applied at age 50, with a background 10-year CAD risk of 5% (odds 1: 19), the *OAPR* = (2.4 : 19) ≈ 1: 8, i.e., false positives outnumber true positives by around eight to one. Reducing the cut-off to reduce the *FPR* to 1% reduces the *DR* to 3%, with 97% of cases missed. Retaining a 5% *FPR* but applying the test in a population with 2% CAD incidence over the same period (background odds 1: 55), e.g., at around 40 years of age, yields an *OAPR* of 1: 23, with false positives outnumbering true positives by just over twenty to one.

**Figure 2.**
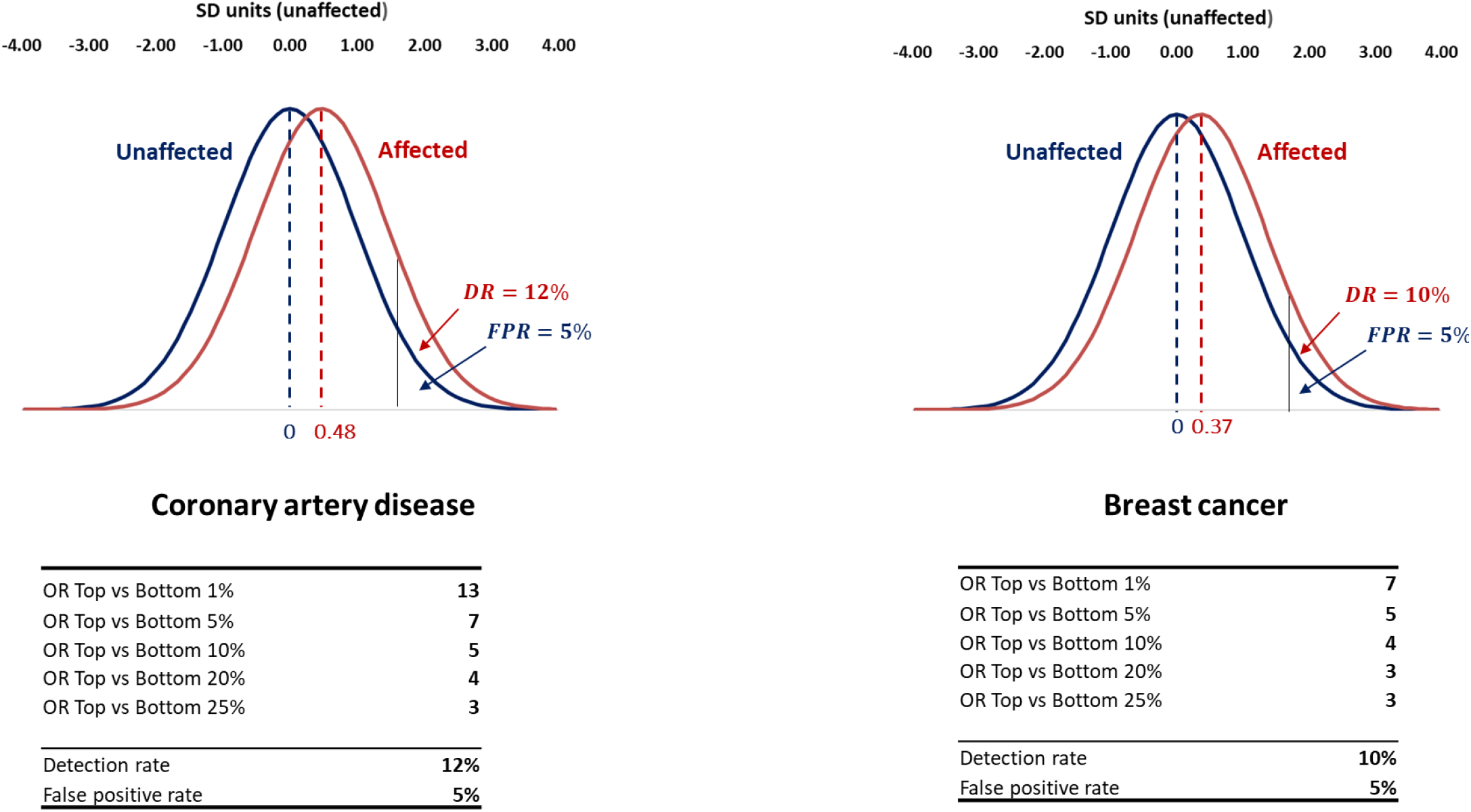
Relative polygenic risk score distributions among those later affected or not by CAD (left panel) and breast cancer (right panel). The mean value of the polygenic risk score distribution those later affected is shifted 0.48 standard deviation units to the right of the mean of the distribution for those who remain unaffected in the case of CAD and 0.37 standard deviation units to the right in the case of breast cancer. The corresponding *DR*_5_ values and odds ratios for comparisons of the top and bottom 1%. 5%, 10%, 20% and 25% of the unaffected polygenic risk score distribution are shown below each plot.

#### Breast cancer

The median *DR*_5_ from performance metrics reported for 108 polygenic risk scores for breast cancer was 10% [9 − 12] (90% of cases missed **Figure 2**). This corresponds to an *LR* of 2. Applied at age 50, with a background 10-year breast cancer risk of about 2.5% (odds 1: 41), the *OAPR* = 1: 21. Applying the polygenic risk score as a test at age 40, when the background 10-year odds is 1: 64 yields an *OAPR* of 1: 32, false positives outnumbering true positives by just over thirty to one.

### Performance of polygenic risk scores in individual risk prediction

The overlap in polygenic score distributions derived from the metrics in the PGS Catalog enables calculation of the *LR* for an individual which, together with the background odds of the disorder for the population, can be used to calculate the odds of becoming affected for that *individual* (see **Extended Methods**)^22^.

#### Coronary artery disease

The odds of developing CAD in the next 10-years were 1: 54, 1: 29, 1: 15, and 1: 8 with a polygenic risk score at the 2.5*th*, 25*th*, 75*th* and 97.5*th* centile respectively at age 50 (when the background odds is 1: 19) (**Figure 3**); and 1: 157, 1: 85, 1: 45, and 1: 24 respectively at age 40 (when the background odds is 1: 55).

**Figure 3.**
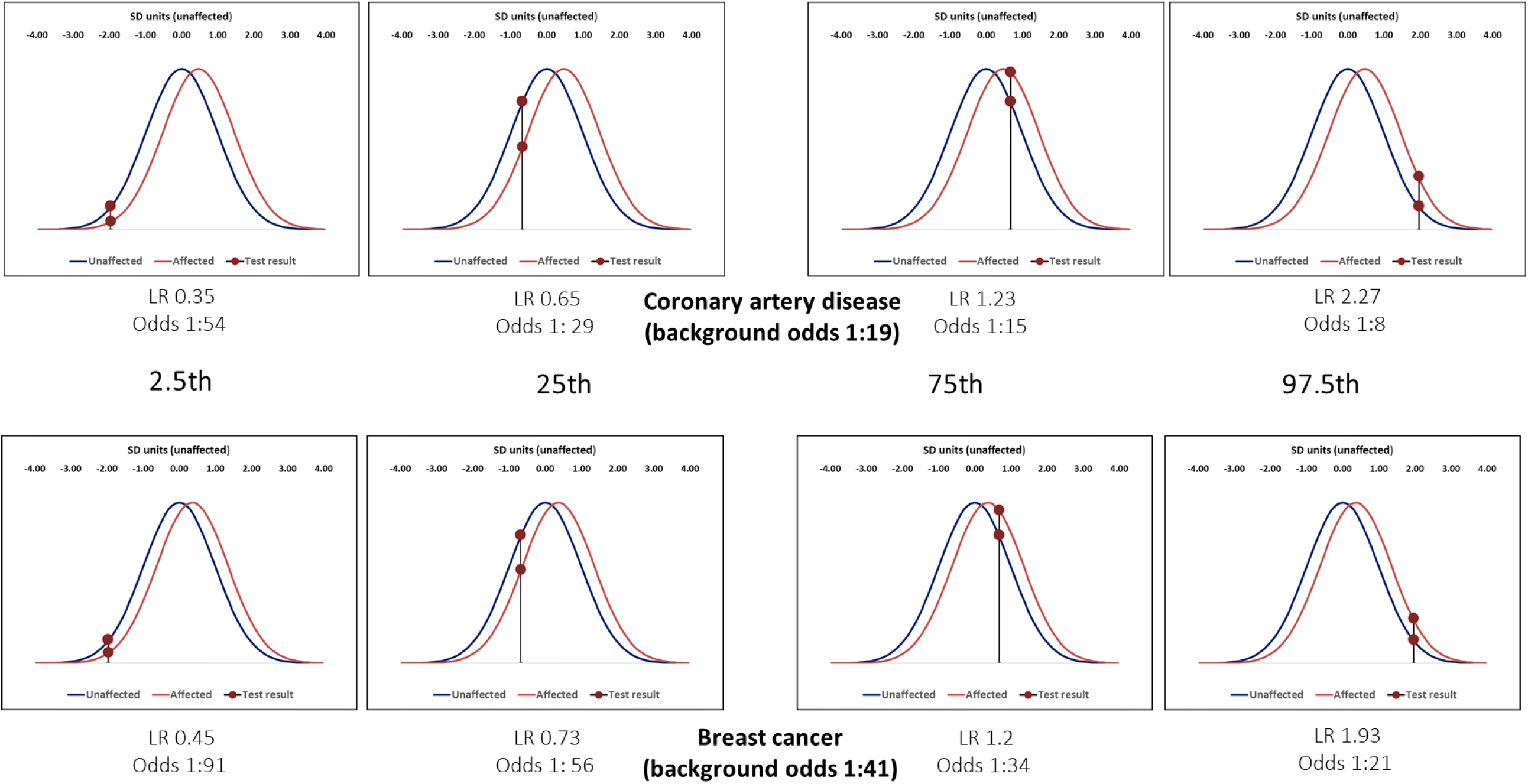
Likelihood ratios and 10-year odds of CAD (top panel) and breast cancer (bottom panel) for 50-year olds with a polygenic risk score result corresponding to the 2.5*th*, 25*th*, 75*th* and 97.5*th* centiles of the corresponding distribution.

#### Breast cancer

The average 10-year odds of breast cancer is 1: 41 for a woman aged 50, and 1: 64 for a woman aged 40. The corresponding odds of being affected were 1: 91, 1: 56, 1: 34, and 1: 21 respectively at age 50 (**Figure 3)**, and 1: 142, 1: 88, 1: 53, and 1: 33 at age 40, for a woman with a polygenic risk score at the 2.5*th*, 25*th*, 75*th* and 97.5*th* centile respectively.

### Performance of polygenic risk scores in risk stratification

#### Coronary artery disease

The left panel in **Figure 4** shows the overlapping distributions for affected and unaffected individuals applied to a hypothetical cohort of 100,000 50-year-old men with stratification into polygenic risk score quintile groups. The 10-year odds of CAD are reduced from the average of 1: 19 for all men to 1: 41 for those in the lowest quintile group and increased to 1: 11 in the highest quintile group. In **Figure 5**, focusing on those at the highest risk, the 10-year odds was 1: 7 for the upper 2.5% of the polygenic risk score distribution for CAD, but this group contributed only 7% of cases (**Figure 5**).

**Figure 4.**
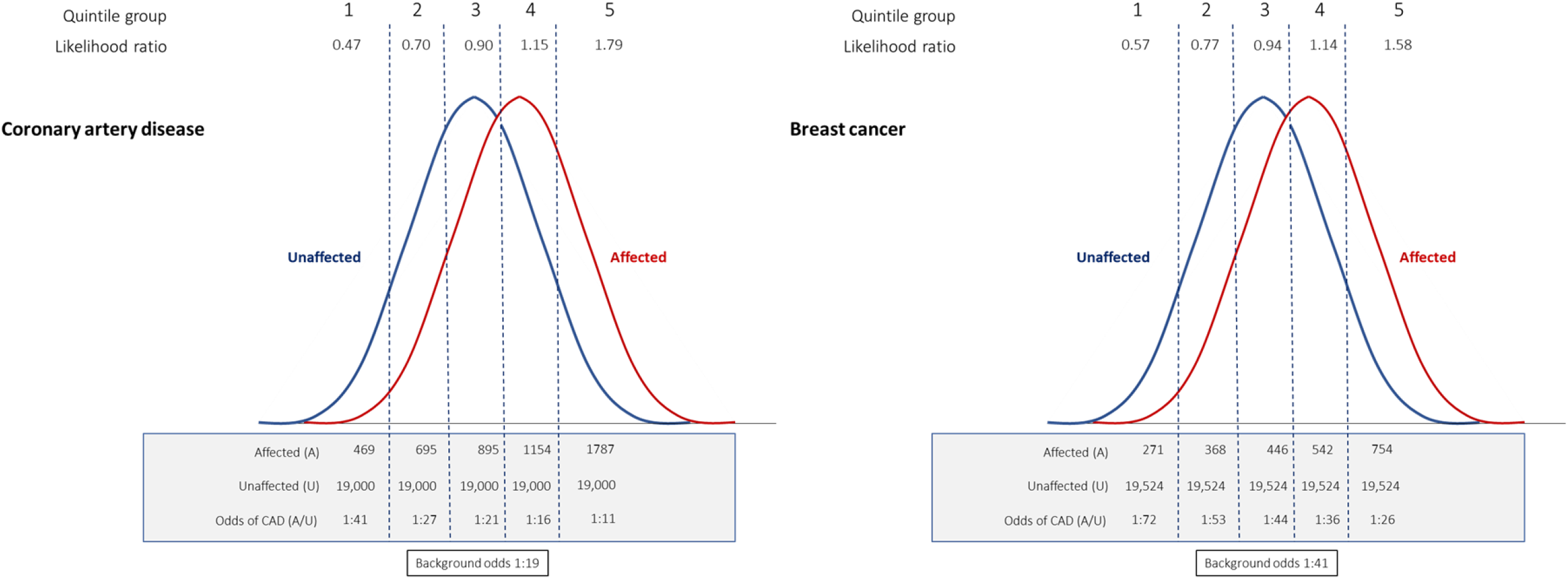
Likelihood ratios, odds and the number of affected and unaffected individuals for each quintile group in a hypothetical population of 100,000 individuals with a background 10-year odds of CAD of 1:19 (left panel) and women with a 10-year odds of breast cancer of 1: 41 (right panel).

**Figure 5.**
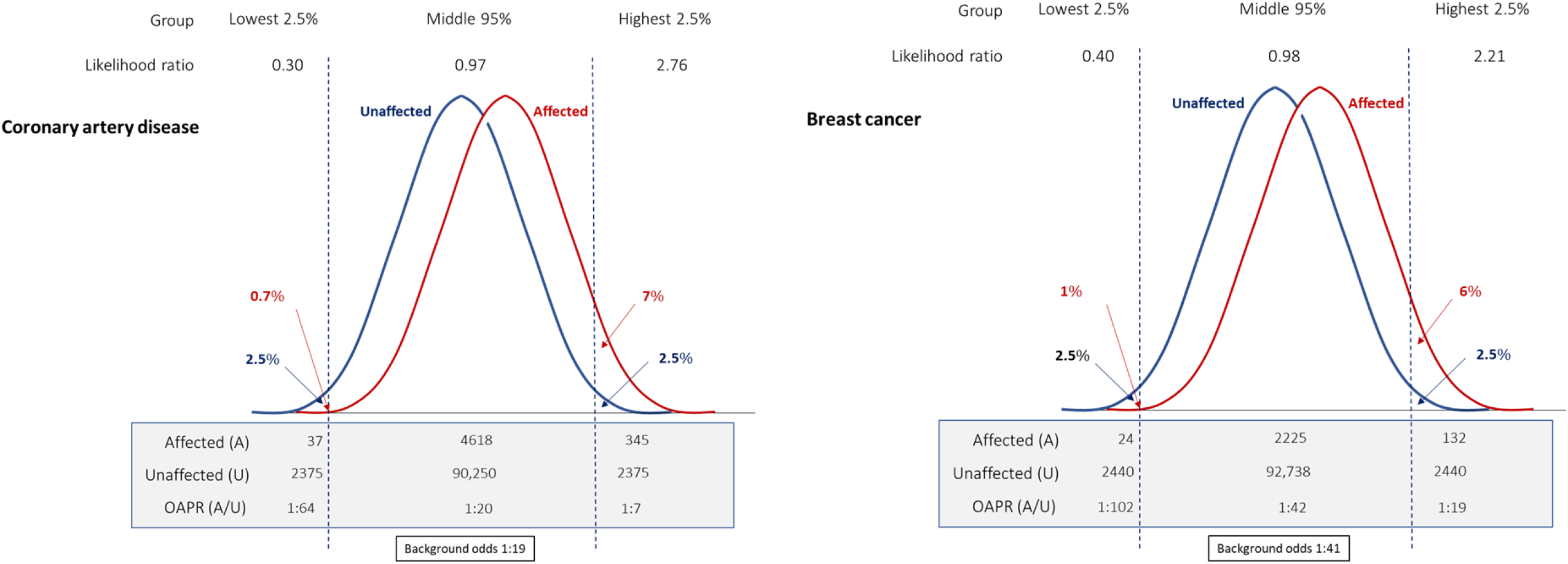
As figure 4 but comparing highest and lowest 2.5% of the unaffeted polygenic risk score distributions for CAD (left panel) and breast cancer (right panel).

#### Breast cancer

In the right-hand panel of **Figure 4**, we applied the same approach to a hypothetical cohort of 50-year-old women with a background 10-year odds of breast cancer of 1: 41. Odds reduce by about half to 1: 72 for those in the lowest quintile and almost double to about 1: 26 for those in the highest quintile of the polygenic risk score distribution. For the highest 2.5% of the polygenic risk score distribution in the right-hand panel of **Figure 5**, odds are increased to 1: 19, but the latter group only accounts for 6% of breast cancer cases.

### Screening using polygenic risk scores together with conventional risk factors or tests

#### i) CAD

It has been proposed that adding polygenic risk scores to conventional risk factors (e.g., blood pressure and LDL-cholesterol), would usefully improve CAD and stroke screening to indicate who should be offered a statin prescription for primary prevention. The **Table** shows the results from Sun *et al*.^23^, applied in a hypothetical cohort of 100,000 40-year old individuals with a risk factor profile representative of the English population and a background 10-year CAD and stroke risk of 8%. A conventional, multi-risk factor model incorporating age and using a 10% 10-year risk cut off, detected 60% of those later affected by CAD or stroke at a 24% *FPR* (*DR*_24_ = 60%). The addition of polygenic risk scores for CAD and stroke to the model (PGS Catalog identifiers PGS000018 and PGS000039 respectively) led to the detection of 61% of those affected for a 23% *FPR* (*DR*_23_ = 61%). Assuming a 10-year risk cut-off of 10% for prescribing statins^24^, 100% adherence, and adopting the assumption of Sun *et al*. that statins reduce the risk of CAD and stroke by 20%^23^, 974 events would be prevented using a model based on conventional risk factors *and* polygenic risk scores instead of 957 using a conventional risk factor model with no genetic information, a gain of 17 cases. This gives a number needed-to-genotype to prevent one additional event of 5882 (**Table**). Sun *et al*. also estimated that 1029 CAD and stroke events would be prevented using a hybrid model, where conventional risk factor assessment is followed by polygenic risk scores only for those at intermediate (5 − 10%) 10-year risk. However, replacing this more complicated model with one where the whole cohort receives statins would prevent 1600 cardiovascular events using the same assumptions (**Supplementary Tables**). Since age is the major determinant of CAD and stroke risk, age alone performs about as well as muti factor risk models that include age^25^. Given the rarity of CAD and stroke events below 50, using an age cut-off of 50 instead of 40 would prevent almost as many events but with many fewer false positives^26^.

**Table.**
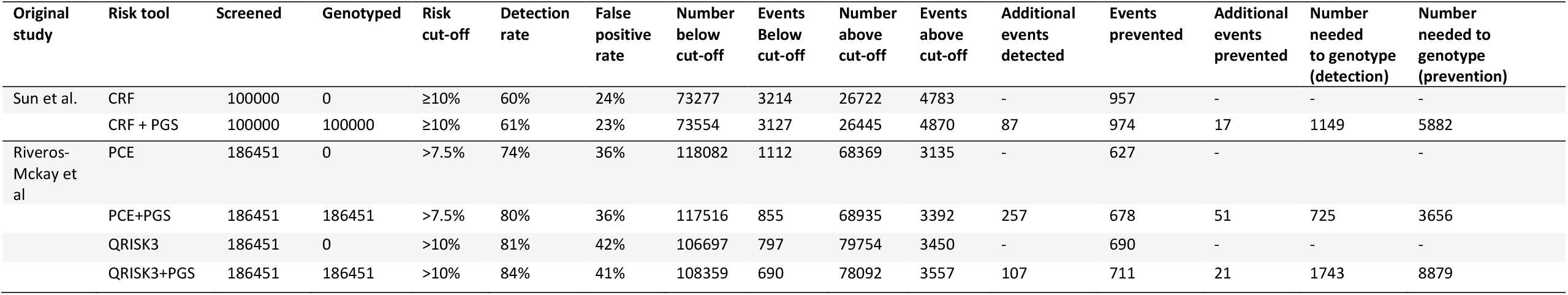
Effect of adding a polygenic risk score to non-genetic risk factors in prediction of coronary artery disease (CAD) and stroke. The values are based on data reported by Sun *et al*. (Ref 23) and Riveros-Mckay *et al*. (Ref 5). Both studies utilised data from UK Biobank. Sun *et al*. developed a conventional risk factor model (CRF) and examined the effect of adding polygenic risk scores for CAD (PGS000018) and stroke (PGS000039) on the prediction of subsequent CAD and stroke events. They used a 10-year risk cut-off of 10% for offering statin treatment. Riveros-McKay *et al*. modelled screening performance in 186,451 participants based on either the Pooled Cohort Equation (PCE) using a 7.5% 10-year risk cut-off, or QRISK3 using a 10% risk cut-off for statin prescription. The data on events reported by Riveros-Mackay *et al*. were for CAD alone rather than CAD and stroke. Calculations assume that all those exceeding the specified risk cut-off receive a statin, 100% adherence and that statin treatment produces a 20% relative risk reduction. Number needed to genotype refers to the number of individuals that need to be genotyped (and have a polygenic risk score calculated) to detect or prevent one additional cardiovascular event.

Similar results were obtained using data reported by Riveros-McKay et al.^5^. These authors also investigated the extent to which the addition of a polygenic risk score to conventional risk factors improves the identification of UK Biobank participants eligible to receive statins because their 10-year risk of CAD and stroke exceeds the cut-offs used in UK or US primary prevention guidelines. Deriving the appropriate metrics from their data (**Table and Supplementary Tables**) clarifies the effect of adding information from a polygenic risk score for CAD. Using a 10-year risk cut-off of 10% for initiating statins, the QRISK3 model based on conventional risk factors including age detected 81% of cases at a 42% *FPR* (*DR*_42_ = 81%). The addition of a polygenic risk score to the model detected 84% of cases for a 41% *FPR* (*DR*_41_ = 84%). Using the authors’ assumption that statins reduce CAD and stroke events by 20%, the number needed-to-genotype to prevent one additional event based on this study is 8879 (**Table**).

#### ii) Breast cancer

It has also been proposed that polygenic risk scores are used to prioritise the use of established screening tests for cancer^3^. One suggestion is that younger women should undergo mammographic screening if their breast cancer risk determined using a polygenic risk score exceeds that of an average 50-year-old, the age beyond which mammography is offered to all women. **Figure 5** shows that 40-year-old women at or above the unaffected 97.5*th* centile of a breast cancer polygenic risk score distribution have an odds of breast cancer of 1: 19, higher than the average 10-year odds at age 50 of 1: 41 ^27^. **Figure 6** shows that using the breast cancer polygenic risk score (PGS Catalog identifier PGS000004) as a stage 1 screen in 100,000 40-year-old women, applying the unaffected 97.5*th* centile as a cut-off, would result in 2570 women with a ‘high-risk’ polygenic score being offered mammography of whom 108 would be affected and 2462 unaffected (*OAPR* 1: 23). Assuming 100% uptake and a *DR*_8_ of 75%^28^, mammography would then correctly identify 81 of the 108 affected individuals but miss 27 breast cancer cases. However, 1430 breast cancer cases (over ten times as many) are estimated to occur among the 97 430 40-year-old women with a polygenic risk score below the unaffected 97.5*th* centile who would not be offered mammography.

**Figure 6.**
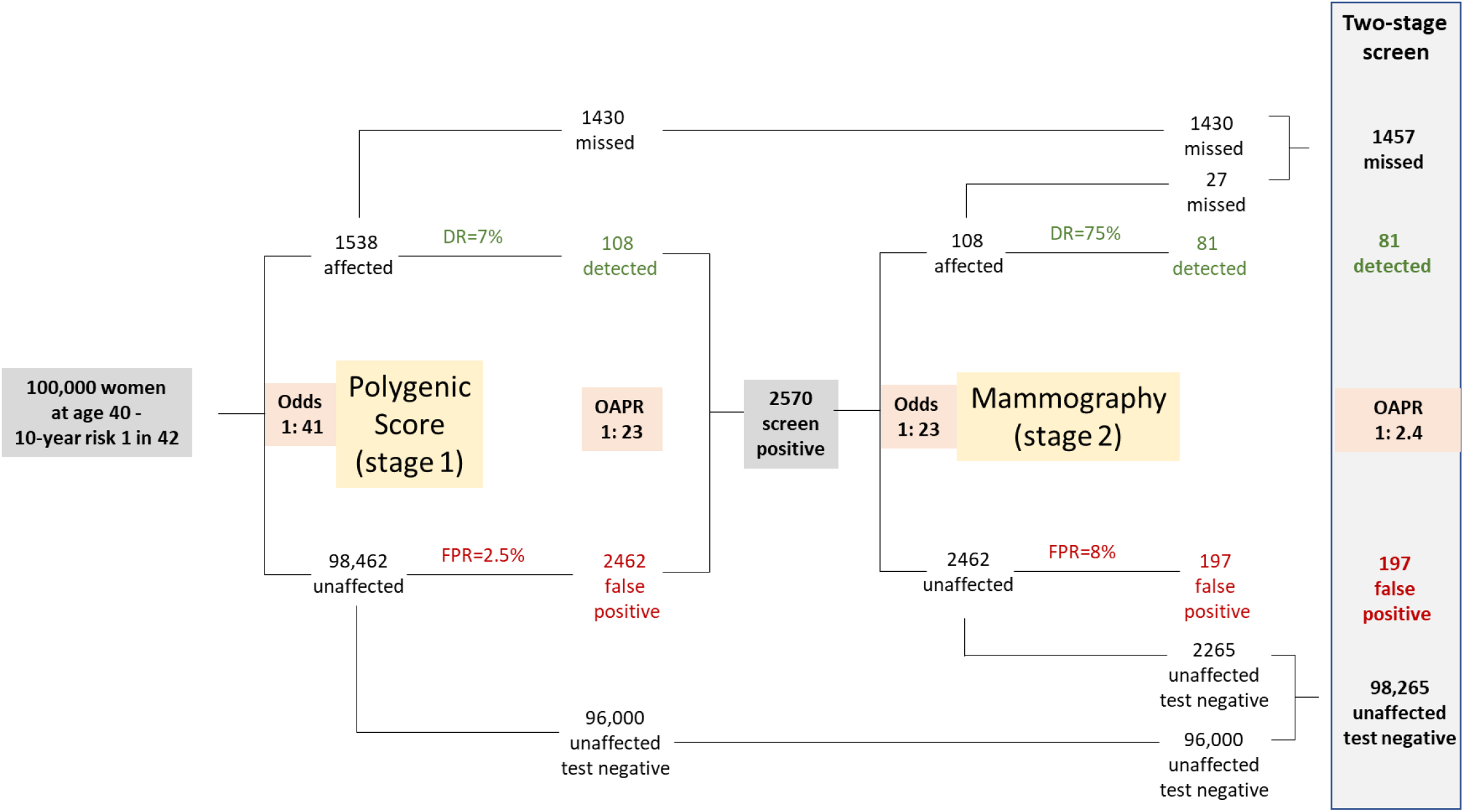
The estimated number of breast cancer cases detected and missed, the number of false positives, and the number of additional mammograms for a two-stage screening test using a polygenic risk score PGS000004 with a cut-off at the unaffected 97.5^th^ centile. Estimates are based on on a hypothetical cohort of 100,000 40-year old women with a background 10-year odds of breast cancer of 1: 41. Performance of mammography in the detection of breast cancer uses estimates from JAMA. 2005; 293:1245

## Discussion

Our results show the poor performance of polygenic risk scores in population screening, individual disease prediction, and population risk stratification. This is not obvious from the metrics reported in the PGS Catalog but is clear using the appropriate metrics employed in this study. Our conclusion is consistent with that of other authors,^17,18,29^ but is insufficiently recognised. The findings are relevant to consumers, patients, doctors, those involved in preventative medicine and public health, as well as funders and policy makers.

Polygenic risk score distributions overlap substantially for all conditions studied and this extensive overlap constrains their performance in each of their intended applications, whether used alone or in conjunction with conventional risk factors or screening tests. For instance, achieving a clinically useful performance in population screening, such as an 80% detection rate for a 5% false positive rate (*DR*_5_ = 80%), requires an *OR* per-*SD* of 12 or higher (compared to the median observed value of 1.31) or an *AUC* of 0.96 (compared to the median observed value of 0.65). Only 11.4% of *AUC* values in the PGS Catalog exceeded 0.8 which equates to a *DR*_5_ of 32%, with most of these resulting from large effect variants at the HLA locus in a few autoimmune diseases (**Figure 1** and **Supplementary Tables**).

Where a risk factor displays a monotonic relationship with disease risk^30^, more cases arise among the majority with near average risk factor values than among the few with more extreme values – the ‘prevention paradox’ ^31 32^. In this respect, polygenic risk scores are similar to certain non-genetic risk factors such as blood pressure and LDL-cholesterol which, though causal, are poor predictors of CAD^17,33^. That the performance of polygenic risk scores in the prediction of CAD is sometimes compared favourably to that of blood pressure and cholesterol^23^ is simply to benchmark one poor predictor against another.

Where there are safe and inexpensive preventative interventions (e.g., statins and blood pressure lowering drugs for prevention of CAD and stroke) there is greater public health benefit in broadening rather than limiting eligibility for such interventions^34^. In the prevention of CAD and stroke, this has been achieved *de facto* by the progressive lowering of the 10-year risk cut-off for statin prescription in primary prevention. The cut-off was reduced from a 30% 10-year risk of CAD in the UK in 1997^35^, down to 10% 10-year risk of CAD or stroke in the UK from 2016^24^ and 7.5% in the US from 2019^36^. The reduction in the risk cut-off was enabled by falling drug acquisition cost through patent expiry, and by accumulating evidence on long-term safety. Eligibility could be extended yet further and also simplified by using age alone to guide statin prescription for primary prevention with the prevention of many more CAD and stroke cases^25^. By contrast, retaining the same 10-year risk cut-off and adding polygenic risk score information to conventional risk factor models has a much weaker impact. Using recently reported data^5,23^, we show several thousand individuals need to be genotyped and a polygenic risk score calculated to prevent one additional vascular event.

Identifying a minority of individuals at very high risk (using genetics or other means) may be justified if a preventative intervention is costly, resource limited, or has significant harms^37^. However, as we demonstrated using breast cancer as an example, identifying those at high risk requires testing in all and, aside from missing the many more cases among those at average risk, generates many false positives. This could have substantial downstream resource implications for healthcare systems if, for example, genetic risk stratification were to be followed by a confirmatory screening test, such as mammography for breast cancer. In this case, it would be more sensible to simply reduce the age cut-off for mammography for all women without determining their polygenic risk score.

The enthusiasm surrounding polygenic risk scores may have been encouraged by pressure on academia to demonstrate a tangible health impact after decades of research investment in human genomics and by commercial opportunity. Unrealistic expectations have probably been raised by use of the wrong metrics. Publications on polygenic risk scores often illustrate comparisons between mutually exclusive groups, e.g., those in opposite tails of a polygenic score distribution^38^. This is relevant in aetiological studies but is not relevant in screening. As shown in **Figure 1**, seemingly impressive odds ratios of 13, 7, 5, 4 or 3, for comparisons of the top vs. the bottom 1%, 5%, 10%, 20% and 25% of the polygenic risk score distribution respectively, all equate to a *DR*_5_ of only 12%. What is relevant in screening is the risk of an event in a group compared to that of the whole population, which is what calculation of the *DR* for a specified *FPR* achieves.

Our findings are relevant both to commercial providers of genetic tests and to researchers working on polygenic risk scores. Commercial providers could communicate individual test results to customers with greater clarity and relevance to performance in disease prediction, e.g., by presenting the overlapping distributions of polygenic risk scores among those later affected and unaffected and by presenting an absolute measure of risk for an individual or group, which requires additional information on population average risk at a particular age over a specified time. In tandem, as already suggested^39^, policy makers may wish to consider tighter regulation of commercial genetic tests based on polygenic risk scores, with a focus on clinical not just assay performance, to protect the public from unrealistic expectations and already stretched public health systems from becoming overburdened by the management of false positive results. Researchers reporting studies on polygenic risk scores should present as a minimum: 1) the mean and *SD* of polygenic risk score values among later affected and unaffected individuals; 2) the overlap in their distributions; 3) the relevant performance metrics such as the *DR* for a specified *FPR*, such as the *DR*_5_, avoiding the need to calculate this indirectly^20^; and 4) the performance of polygenic risk scores with and without the inclusion of other variables so that users can judge the incremental benefit provided by the polygenic risk score itself.

Although the current analysis shows the poor performance of polygenic risk scores in screening, prediction, and risk stratification, they may find use in other situations. For example, polygenic scores may explain the variable penetrance of rare mutations in monogenic diseases, e.g., hypertrophic cardiomyopathy or familial hypercholesterolaemia, and be employed to aid case detection. There are also other predictive applications of genotyping, e.g., in pharmacogenetic testing, to optimise efficacy and safety of medicines. Genotyping may also be of value in blood and tissue matching. Because genetic variation is transmitted from parents to offspring through a randomised process (like treatment allocation in a clinical trial), and is unaltered by disease, an important translational application arising from genomic discoveries is likely to be in providing evidence on disease causation and targets for pharmaceutical intervention^40^.

In conclusion, use of the appropriate metrics has demonstrated the poor performance of polygenic risk scores in population screening, individual risk prediction, and population risk stratification. By virtue of its wide scope, the current study may help to resolve the debate on the value of polygenic risk scores and avoid unjustified expectations about their role in preventing disease.

## Data Availability

All data produced in the present work are contained in the manuscript

## Acknowledgement of funding

Supported by the UCL British Heart Foundation Accelerator (AA/18/6/34223), the UCL NIHR Biomedical Research Centre, and the UKRI/NIHR funded Multimorbidity Mechanism and Therapeutics Research Collaborative (MR/V033867/1). JG is a UCL British Heart Foundation PhD candidate (FS/17/70/33482). ADH and HH are NIHR Senior Investigators.

## Data sharing

The data used in this manuscript are in the Supplementary Tables.

