## Extended Methods for "Performance of polygenic risk scores in screening, prediction, and risk stratification"

**Hingorani AD., *et al*.**

Our calculations assume polygenic scores exhibit a Gaussian distribution in a population; the proportional difference in disease risk is the same for any given absolute difference in polygenic score value, from any starting level (a log-linear relationship); and that polygenic score distributions have the same standard deviation in affected and unaffected individuals. The first assumption has both empirical and theoretical support according to the Central Limit Theorem; the second follows from the way that polygenic scores are constructed, and their performance reported (as an $OR$ or $HR$ per $SD$ increment in the polygenic score value); and the third assumption also has substantial empirical support, e.g., from the colorectal cancer scores in UK Biobank, as reported in Supplementary table 3 of *Lambert et al*.

*Deriving $DR_5$ from $HR$ or $OR$ per-SD*

The detection rate ($DR$) for a given false positive rate can be derived from the $OR$ or $HR$ per-$SD$ as follows. Consider the Gaussian frequency distributions of polygenic risk scores among those unaffected ($U$) and those who become affected by disease ($A$). The mean polygenic score value among those unaffected is $\mu_U$ and among those affected is $\mu_A$, and both distributions have the same $SD$ ($\sigma$)

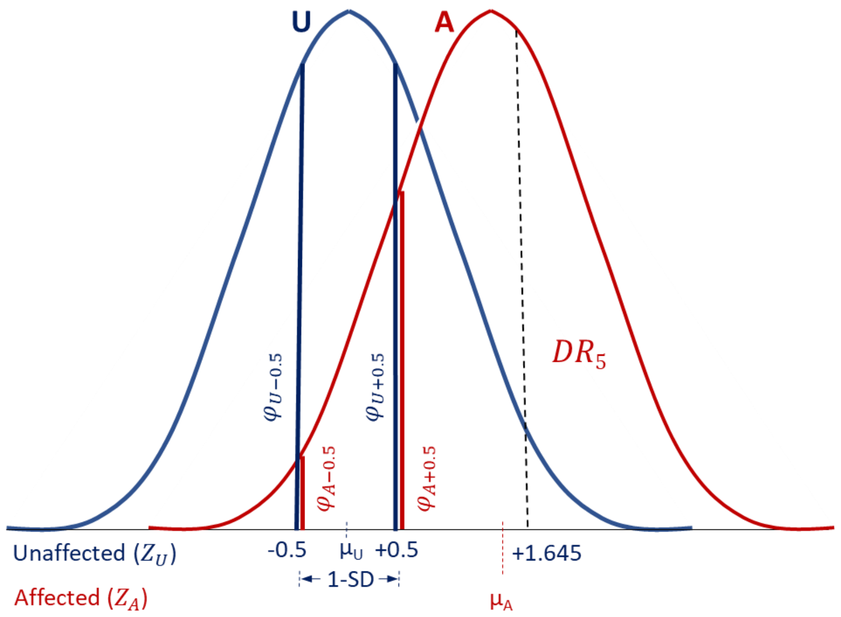


The $OR$ per-$SD$ can be considered to be equal to the ratios of the probability density function values $\varphi$, for affected ($\varphi_A$) vs. unaffected individuals ($\varphi_U$) at Z-score values for the distribution among unaffecteds (i.e. $Z_U$ values) that are 1-$SD$ apart, for example corresponding to values at $+0.5\ SD$ and $-0.5SD$. In the figure, the values are shown by vertical red and blue, lines respectively. Thus,

$$OR_{SD} = \frac{\left(\varphi_A/\varphi_U\right)_{+0.5}}{\left(\varphi_A/\varphi_U\right)_{-0.5}}$$

Where $x$ is the value of the variate, the probability density function $\varphi$ is given by:

$$\varphi(\mu, \sigma, x) = \frac{1}{\sigma\sqrt{2\pi}} e^{-\frac{1}{2}\left(\frac{x-\mu}{\sigma}\right)^2}$$

For the distribution in the unaffected, $\mu$ = 0, $\sigma$ =1

$$\varphi U(0,1,x) = \frac{1}{1\sqrt{2\pi}} e^{-\frac{1}{2}\left(\frac{x}{1}\right)^2}$$

$$\varphi U(0,1,-0.5) = \frac{1}{1\sqrt{2\pi}} e^{-\frac{1}{2}\left(\frac{-0.5}{1}\right)^2}$$

$$\varphi U(0,1,+0.5) = \frac{1}{1\sqrt{2\pi}} e^{-\frac{1}{2}\left(\frac{+0.5}{1}\right)^2}$$

Since $\varphi_{U+0.5}$= $\varphi_{U-0.5}$,

$$OR_{SD} = \frac{(\varphi_{A+0.5})}{(\varphi_{A-0.5})}$$

For the distribution in the affected, $\mu$ = $\mu_A$, $\sigma$ =1

$$\varphi A(\mu_A, \sigma, -0.5) = \frac{1}{1\sqrt{2\pi}} e^{-\frac{1}{2}\left(\frac{-\mu_A-0.5}{1}\right)^2}$$

$$\varphi A(\mu_A, \sigma, +0.5) = \frac{1}{1\sqrt{2\pi}} e^{-\frac{1}{2}\left(\frac{0.5-\mu_A}{1}\right)^2}$$

Thus,

$$OR_{SD} = \frac{\frac{1}{1\sqrt{2\pi}} e^{-\frac{1}{2}\left(\frac{0.5-\mu_A}{1}\right)^2}}{\frac{1}{1\sqrt{2\pi}} e^{-\frac{1}{2}\left(\frac{-\mu_A-0.5}{1}\right)^2}}$$

Cancelling $\frac{1}{1\sqrt{2\pi}}$,

$$OR_{SD} = \frac{e^{-\frac{1}{2}(0.5-\mu_A)^2}}{e^{-\frac{1}{2}(-\mu_A-0.5\,)^2}}$$

$$OR_{SD} = e^{[-\frac{1}{2}(0.5-\mu_A)^2+\frac{1}{2}(-\mu_A-0.5\,)^2]}$$

$$OR_{SD} = e^{-\frac{1}{2}[\,(0.5-\mu_A)^2-\,(-\mu_A-0.5\,)^2]}$$

$$OR_{SD} = e^{-\frac{1}{2}[-2\mu_A]}$$

$$OR_{SD} = e^{\mu_A}$$

$$\mu_A = \ln OR_{SD}$$

Also from the figure,

$$DR_5 = 1 - \emptyset(1.645 - \mu_A)$$

where $\emptyset$ corresponds to the cumulative distribution function ($CDF$) corresponding to the area under the normal distribution among affecteds at a $Z$-value of $1.645 - \mu_A$. (The $1.645$ is the $95^{th}$ centile of the distribution among unaffecteds, which is the threshold for a $5\%$ false positive rate). Because of the symmetry of the normal distribution,

$$DR_5 = \emptyset(\mu_A - 1.645)$$

Thus, from equation 2,

$$DR_5 = \emptyset(\ln OR_{SD} - 1.645)$$

Consider, as an example, an $OR_{SD} = 5$.
In this example, $\ln OR_{SD} = \mu_A = 1.609$.
Thus, from equation 3,

$$DR_5 = \emptyset(1.609 - 1.645)$$
$$DR_5 = \emptyset(-0.03556)$$
$$DR_5 = 0.485816$$

Thus an $OR$ per-$SD$ of $5$ corresponds to a $49\%$ detection rate for a $5\%$ false positive rate.

*Estimating $DR_5$ from AUC and C-index*

The following derivation is adopted from Wald and Bestwick (Ref 21).

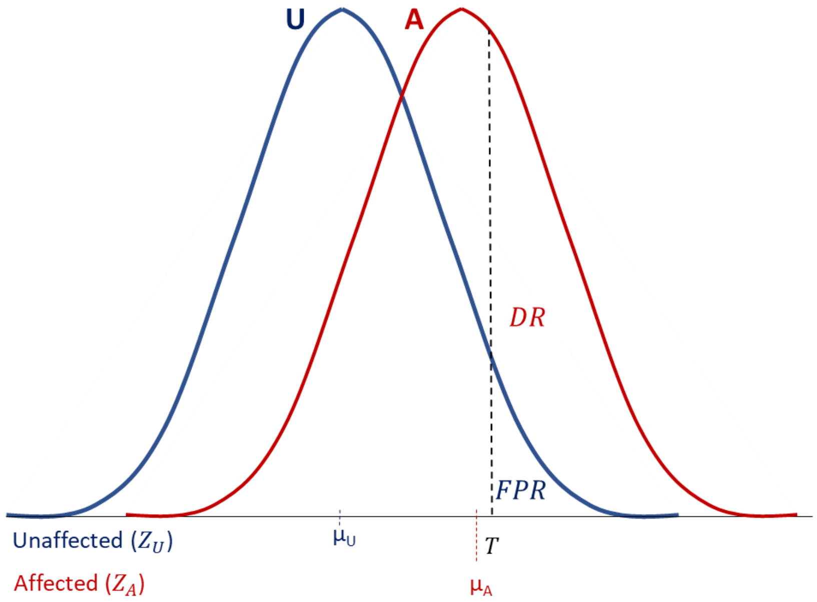


The $DR$ at a cut-point value T, is given by

$$DR = 1 - \emptyset\left(\frac{T - \mu_A}{\sigma_A}\right)$$

Because of the symmetry of the normal distribution,

$$DR = \emptyset\left(\frac{\mu_A - T}{\sigma_A}\right)$$

By the same argument,

$$FPR = \emptyset\left(\frac{\mu_U - T}{\sigma_U}\right)$$

The $Z$-score corresponding to the $DR$ expressed in terms of the affected distribution is:

$$\emptyset^{-1}(DR) = \frac{\mu_A - T}{\sigma_A}$$

The $Z$-score corresponding to the FPR expressed in terms of the unaffected distribution is:

$$\emptyset^{-1}(FPR) = \frac{\mu_U - T}{\sigma_U}$$

Writing both equations in terms of $T$

$$T = \mu_A - \sigma_A \emptyset^{-1}(DR)$$

$$T = \mu_U - \sigma_U \emptyset^{-1}(FPR)$$

Thus,

$$\mu_A - \sigma_A \emptyset^{-1}(DR) = \mu_U - \sigma_U \emptyset^{-1}(FPR)$$

Making $DR$ the subject,

$$\sigma_A \emptyset^{-1}(DR) = \mu_A - \mu_U + \sigma_U \emptyset^{-1}(FPR)$$

$$\emptyset^{-1}(DR) = \frac{\mu_A - \mu_U + \sigma_U \emptyset^{-1}(FPR)}{\sigma_A}$$

$$DR = \emptyset\left(\frac{\mu_A - \mu_U}{\sigma_A} + \frac{\sigma_U \emptyset^{-1}(FPR)}{\sigma_A}\right)$$

Considering the standard normal distribution and assuming equal standard deviation in unaffected and affected groups, and $\mu_U = 0$.

$$DR = \emptyset\left(\frac{\mu_A}{1} + \frac{1 \cdot \emptyset^{-1}(FPR)}{1}\right)$$

$$DR = \emptyset\big(\mu_A + \emptyset^{-1}(FPR)\big)$$

The area under the ROC curve (*AUC*), or equivalently the $C$-index, is the probability that an affected individual drawn at random ($A$) has a higher polygenic risk score than an unaffected individual drawn at random ($U$) i.e.

$$P(A > U) = P(A - U > 0)$$

The $AUC$ is the therefore the CDF for the distribution of differences (the variances sum). Thus,

$$AUC = \emptyset\left(\frac{\mu_A - \mu_U}{\sqrt{\sigma_A^2 + \sigma_U^2}}\right)$$

$$\emptyset^{-1}(AUC) = \frac{\mu_A - \mu_U}{\sqrt{\sigma_A^2 + \sigma_U^2}}$$

Given that for the standard normal distribution $\mu_U = 0$ and if we assume equal standard deviation for the distributions for affected and unaffected individuals, $\sigma_U = \sigma_A = 1$,

$$\emptyset^{-1}(AUC) = \frac{\mu_A}{\sqrt{2}}$$

For example, if the $AUC$ is $0.7$,

$$\emptyset^{-1}(0.7) = 0.524$$

Thus,

$$\frac{\mu_A}{\sqrt{2}} = 0.524$$

$$\mu_A = 0.524 \cdot \sqrt{2}$$

$$\mu_A = 0.742$$

Since,

$$DR = \emptyset\left(\mu_A + \emptyset^{-1}(FPR)\right)$$

the $DR_5$ corresponding to an $AUC$ of $0.7$ is,

$$DR_5 = \emptyset\left(0.742 + \emptyset^{-1}(0.05)\right)$$

$$DR_5 = \emptyset(0.742 + (-1.645)$$

$$DR_5 = \emptyset(-0.903)$$

$$DR_5 = 0.183$$

Thus an $AUC$ of $0.7$ corresponds to a detection rate of $18\%$ for a $5\%$ false positive rate.

*Calculating the likelihood ratio and odds of becoming affected given a positive test result ($OAPR$)*

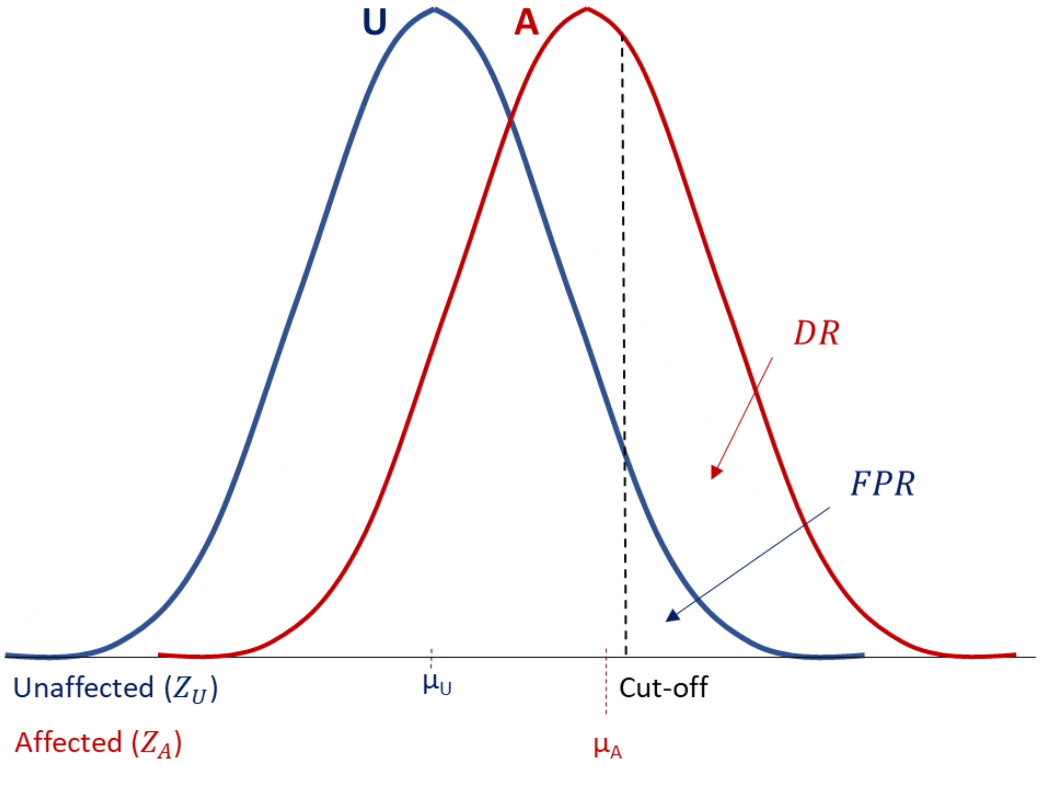


In evaluating the performance of a polygenic risk score as a screening test, we calculated the likelihood ratio for a positive result (i.e., a polygenic risk score at or above a pre-specified cut-off) as the ratio $DR/FPR$. The likelihood ratio for a test cut-off with a $5\%$ $FPR$ is given by $^{DR_5}/_{5\%}$, where $DR_5$ is expressed as a percentage. The $OAPR$ is calculated by multiplying the background odds of disease by the likelihood ratio for a positive test result. For example, if the background odds of disease in the population is $1{:}9$ and the $DR_5$ is $15\%$, the likelihood ratio is $15/5 = 3$ and the $OAPR = 3{:}9$ or $1{:}3$.

*Calculating the likelihood ratio and odds of becoming affected for an individual with a given polygenic score result*

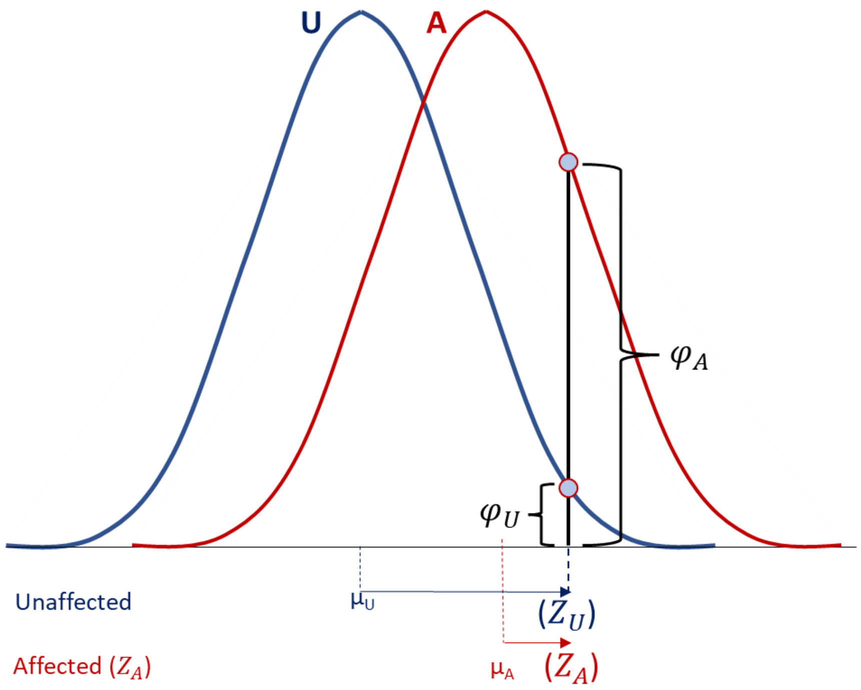


To compute how many times more likely a given polygenic score result is to arise from an affected than unaffected individual, we compared the heights of the standard distribution curves for affected and unaffected individuals at that value (the likelihood ratio; $LR$). In the illustration shown, this is given by the ratio ${}^{\varphi_A}/_{\varphi_U}$.

$$LR = {}^{\varphi_A}/_{\varphi_U}$$

These heights can be calculated using the equation for the Gaussian distribution:

$$LR = \frac{\frac{1}{\sigma\sqrt{2\pi}} e^{-\frac{1}{2}(Z_A)^2}}{\frac{1}{\sigma\sqrt{2\pi}} e^{-\frac{1}{2}(Z_U)^2}}$$

For example, for a polygenic risk score with a performance metric expressed as an $OR_{SD} = 1.61$:

$$\mu_A = \ln OR_{SD} = 0.48$$

A polygenic score at the 75th centile of the distribution for unaffected individuals yields,

$Z_U = 0.67; \varphi_U = 0.32$

$Z_A = 0.2, \varphi_A = 0.39$

$LR = {}^{\varphi_A}/_{\varphi_U} = {}^{0.39}/_{0.32} = 1.23$

If the background odds of disease in the population is $1:9$, an individual whose polygenic score is at the 75th centile of the distribution among unaffecteds has an odds of being affected of $(1.23 \times 1): 9 \approx 1:7$.

*Calculating the likelihood ratio and odds of becoming affected for a particular polygenic risk score group*

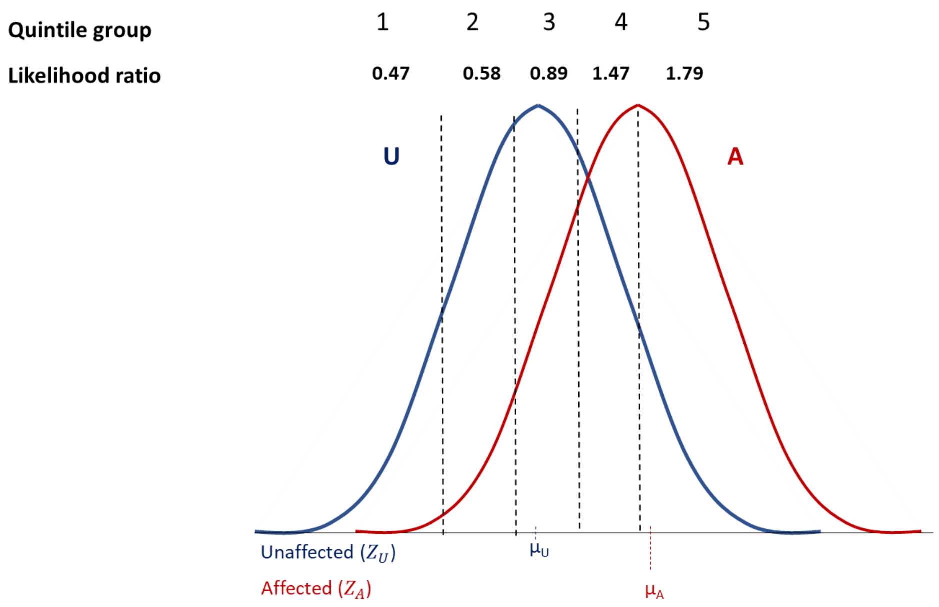


In evaluating polygenic risk scores in risk stratification, we calculated the $LR$ as the ratio of areas under the distributions for affected and unaffected individuals in each polygenic score

quantile (e.g., each fifth of the polygenic score distribution with respect to the unaffected, as shown in the figure). We then multiplied the background odds of disease by the corresponding likelihood ratio to determine the odds of becoming affected for each quantile of the distribution. For example, for individuals in the fourth quintile, the $LR$ is $1{:}\,47$. If the background odds of disease in the population is $1{:}\,9$ the odds of becoming affected for this group is $1.47{:}\,9 \approx 1{:}\,6$.
